# Osteosarcopenia and its impact on falls in older patients in Vietnam

**DOI:** 10.64898/2026.11.25.26369005

**Authors:** Tan Van Nguyen, Mai Tran Thi Bich Duyen, Minh Tung Dao, Thien Vu, Chi L. K. Nguyen, Patrice Ngangue, Mark Woodward, Tu N. Nguyen

## Abstract

**Background and aim:** Osteosarcopenia, the concurrent presentation of osteoporosis and sarcopenia, can increase the risk of physical trauma and functional decline in older adults. This study aimed to investigate the prevalence of osteosarcopenia, its associated factors, and its impact on falls among older hospitalised patients in Vietnam.

**Methods:** This prospective observational cohort study included patients aged 60 years or older admitted to the Department of Geriatric Medicine at Gia Dinh People’s Hospital, Ho Chi Minh City, Vietnam. Participants were followed for 3 months post-discharge to record incident falls. Multivariable logistic regression was applied to examine the factors associated with osteosarcopenia, and the relationship between osteosarcopenia and falls. Results are presented as odd ratios (OR) with 95% confidence intervals (CIs).

**Results:** There were 243 participants (64.6% female). They had a mean age of 75.0 years (SD 8.6 years). The prevalence of osteosarcopenia was 42.0%. Age (OR 1.05, 95% CI 1.01 – 1.09), female sex (OR 9.75, 95% CI 4.57 – 20.82), and underweight (OR 2.48, 95% CI 1.03 – 5.96) were associated with greater odds of having osteosarcopenia. The OR relating osteosarcopenia and falls after 3 months was 2.76 (95% CI 1.20 – 6.37) before adjustment and 2.11 (95% CI 0.82 – 5.39) after accounting for age, sex, rural/urban living and comorbidities.

**Conclusions:** Osteosarcopenia was highly prevalent among older hospitalised adults in Vietnam and was associated with increased odds of falls. Clinicians should consider screening for osteosarcopenia in older adults, particularly among women and those who are underweight.

## Introduction

Osteosarcopenia, the concurrent presentation of sarcopenia and osteoporosis, is a critical geriatric syndrome characterised by the synergistic deterioration of the musculoskeletal unit ^1,2^. Driven by deleterious bone-muscle “cross-talk” via mechanical, inflammatory, and endocrine pathways, structural decline in one tissue accelerates degradation in the other ^3^. With current epidemiological data indicating a prevalence of 21–22% among older populations in Asia and Australia, osteosarcopenia represents a severe and escalating public health burden^4,5^. Clinically, this dual impairment acts as a disease multiplier rather than a simple additive pathology; compared to isolated bone or muscle depletion, osteosarcopenia amplifies the risk of adverse clinical trajectories, precipitating significantly higher incidences of frailty, recurrent falls, hip fractures, and all-cause mortality ^6-8^. In a recent comprehensive meta-analysis of prospective observational studies (with a total of 14429 participants, mean age 70 years, 65% female), osteosarcopenia was associated with approximately 53% increased risk of mortality ^8^.

Research in Vietnam regarding older adults has primarily focused on sarcopenia and osteoporosis as separate conditions. Studies of outpatients in Vietnam reported a sarcopenia prevalence ranging from 41% to 55%, while the prevalence was even higher among older hospitalised patients at 60% ^9-11^. In terms of osteoporosis, a study conducted between 2015 and 2016 involving 1421 women and 652 men aged 50 years and older in the general population of Ho Chi Minh City found that 27% of women and 13% of men had osteoporosis ^12^, while another study conducted in 2025 among 957 adults aged 50 and above in Hue City reported a prevalence of 44% ^13^. There is a lack of studies examining the co-occurrence of these two conditions, both in the general older population and among hospitalised older patients. To address this gap, we conducted this study aimed at providing preliminary insights into the prevalence of osteosarcopenia and its associated factors among older hospitalised patients, and its impact on falls in this population. Our goal is to inform the development of effective strategies for the prevention, diagnosis, and treatment of this condition for older adults in Vietnam.

## Methods

### Study populations

This prospective, observational cohort study was conducted among patients admitted to the Department of Geriatric Medicine of Gia Dinh People’s Hospital in Ho Chi Minh City from November 2021 to May 2022. Consecutive patients aged ≥ 60 years admitted to the Department of Geriatric Medicine during the study period were recruited. The exclusion criteria were: (1) refusal to participate in the study, (2) inability to obtain consent, (3) inability to walk due to any underlying medical condition, (4) having severe acute conditions such as sepsis, hypoglycemic coma, ketoacidosis, severe hepatic failure, acute stroke, acute altered consciousness, or delirium, (5) inability to answer the questionnaire or to perform the instructed movements.

The study was approved by the Ethics Committee of the University of Medicine and Pharmacy at Ho Chi Minh City (Reference Number 284/HDDD-DHYD) and the Ethics Committee of Gia Dinh People’s Hospital (Reference Number 67/NDGD-HDDD). Informed consent was obtained from all participants. This study was conducted in accordance with the Declaration of Helsinki.

### Data collection

Data were collected from patient interviews and medical records. Information obtained from medical records included socio-demographic characteristics (age, sex, rural/urban living), height, weight, medical history, and comorbidities. Comorbidities were documented based on a predefined list, including hypertension, coronary heart disease, heart failure, atrial fibrillation, stroke, dyslipidemia, diabetes, chronic kidney disease, osteoarthritis, gastroesophageal reflux disease (GERD), chronic liver disease, gout, dementia, Parkinson’s disease, chronic lung disease (chronic obstructive pulmonary disease/asthma), and cancer.

Body mass index (BMI) was calculated by dividing weight by height squared (kg/m^2^) and categorised as underweight (<18.5), normal (18.5 -□24.9), or overweight/obese (≥25.0) based on the World Health Organization criteria. Nutritional status was assessed using the Mini Nutritional Assessment Short Form (MNA-SF), and participants with a score ≤7 were classified as having malnutrition, those scoring 8-11 were considered at risk of malnutrition, and scores ≥12 indicated normal nutritional status ^14^. Activities of Daily Living (ADLs) and Instrumental Activities of Daily Living (IADLs) were assessed using questionnaires, and participants with an ADL score <6 were identified as having ADL impairment, whereas those with an IADL score <8 were identified as having IADL impairment ^15^.

Osteosarcopenia was defined as the coexistence of osteoporosis and sarcopenia ^6,16^. Bone mineral density (BMD) and muscle mass were measured using dual X-ray absorptiometry (DXA Hologic) at two locations: the lumbar spine (L2-L4) and the femoral neck. A diagnosis of osteoporosis was confirmed when the T-score at the lumbar spine or femoral neck was ≤ - 2.5 standard deviations for healthy young adults based on the widely used World Health Organization criteria ^17^. Sarcopenia was diagnosed according to the Asian Working Group for Sarcopenia 2019 consensus (Chen et al., 2020) as decreased muscle mass with muscle weakness and/or low walking speed ^18^. The skeletal muscle mass index (SMI) was calculated by the skeletal muscle mass (ASM) of the extremities divided by the square of height in meters (SMI = ASM / height^2^ (kg/m^2^). Decreased muscle mass is defined as a skeletal muscle index SMI <7.0 kg/m^2^ for men and <5.4 kg/m^2^ for women. Handgrip strength was measured using a Jamar hydrostatic dynamometer. The patient sits in a chair with their elbows bent at a 90-degree angle and gradually and continuously squeezes their right and left hands for 3 seconds. The maximum handgrip strength on each side is recorded in kilograms. Muscle weakness was defined as reduced grip strength (<28 kg for men and <18 kg for women). Walking speed was calculated by having the patient walk at their normal speed over a distance of 10 meters. A researcher measured the time taken to walk from 2 meters to 8 meters. Then, the walking speed (v) was calculated using the formula v = s/t, where t is in seconds and s is the distance traveled (6m). Reduced walking speed was defined as v <1 m/s for both sexes.

All participants were followed up for 3 months after discharge, recording incident falls. The information was obtained through phone calls to the participants or their caregivers, and by reviewing participants’ medical records.

### Sample size calculation

Sample size was calculated for the first aim of this study, which was to establish the percentage of people in the target population with osteosarcopenia. Based on previous studies in Asia ^19-21^, we estimated that the prevalence of osteosarcopenia in older patients would be at least 17%. Assuming the usual 5% significance test, to identify an actual prevalence within ±5% with 95% power, the minimum sample size for this study was calculated to be 217 participants.

### Statistical analysis

Continuous variables are presented as means and standard deviation (SD), and categorical variables as frequencies and percentages. Comparisons between participants with and without osteosarcopenia were conducted using the chi-square test or Fisher’s exact test for categorical variables. For continuous variables, their distributions were assessed using Q-Q plots, and Student’s t-tests or Mann-Whitney U tests were applied accordingly to compare participants with and without osteosarcopenia.

Multivariable logistic regression was applied to examine the factors associated with osteosarcopenia. Variables for inclusion in the multivariable logistic regression model were selected based on clinical relevance and univariable analyses (p < 0.20). Potential multicollinearity between these explanatory variables was assessed before model development.

To quantify the association between osteosarcopenia and falls, multivariable logistic regression models were applied, adjusting for age, sex, rural/urban locality, and number of comorbidities. These covariates were selected a priori based on clinical relevance, the available data, and their potential role as confounders of the relationship between osteosarcopenia and the study outcome.

The results are presented as odds ratios (OR) with 95% confidence intervals (CIs). All statistical tests were two-sided and a p value <0.05 was considered to confer statistical significance.

## Results

### Baseline characteristics

A total of 243 participants were included in this study, of whom 102 (42%) were diagnosed with osteosarcopenia (**Table 1**). The number participants with sarcopenia, osteoporosis, and osteosarcopenia among 243 participants is presented in **Figure 1**.

**Table 1.** Participant characteristics.

| Variables | All participants (N=243) | Without osteosarcopenia (N=141) | With osteosarcopenia (N=102) | p-values |
|---|---|---|---|---|
| Age (years) | 74.98 (8.56) | 73.69 (8.71) | 76.76 (8.05) | 0.005 |
| Sex |  |  |  |  |
| Male | 86 (35.4) | 70 (49.6) | 16 (15.7) | <0.001 |
| Female | 157 (64.6) | 71 (50.4) | 86 (84.3) |  |
| Rural/urban living |  |  |  |  |
| Rural | 26 (10.7) | 15 (10.6) | 11 (10.8) | 0.971 |
| Urban | 217 (89.3) | 126 (89.4) | 91 (89.2) |  |
| Body mass index (kg/m<sup>2</sup>) |  |  |  |  |
| <18.50 | 39 (16.0) | 18 (12.8) | 21 (20.6) | 0.048 |
| 18.50 – 24.99 | 169 (69.5) | 97 (68.8) | 72 (70.6) |  |
| ≥ 25.0 | 35 (14.4) | 26 (18.4) | 9 (8.8) |  |
| MNA-SF score |  |  |  | 0.004 |
| ≥ 12 | 54 (22.2) | 42 (29.8) | 12 (11.8) |  |
| 8 – 11 | 143 (58.8) | 74 (52.5) | 69 (67.6) |  |
| ≤7 | 46 (18.9) | 25 (17.7) | 21 (20.6) |  |
| ADL impairment | 82 (33.7) | 43 (30.5) | 39 (38.2) | 0.208 |
| IADL impairment | 189 (77.8) | 106 (75.2) | 83 (81.4) | 0.252 |
| Comorbidities |  |  |  |  |
| Hypertension | 159 (65.4) | 89 (63.1) | 70 (68.6) | 0.373 |
| GERD | 86 (35.4) | 49 (34.8) | 37 (36.3) | 0.806 |
| Dementia | 78 (32.1) | 43 (30.5) | 35 (34.3) | 0.529 |
| Diabetes | 76 (31.3) | 49 (34.8) | 27 (26.5) | 0.169 |
| Coronary heart disease | 68 (28.0) | 32 (22.7) | 36 (35.3) | 0.031 |
| Osteoarthritis | 48 (19.8) | 27 (19.1) | 21 (20.6) | 0.781 |
| Dyslipidemia | 46 (18.9) | 23 (16.3) | 23 (22.5) | 0.221 |
| Chronic lung disease | 35 (14.4) | 22 (15.6) | 13 (12.7) | 0.531 |
| Chronic kidney disease | 31 (12.8) | 20 (14.2) | 11 (10.8) | 0.433 |
| Cancer | 25 (10.3) | 17 (12.1) | 8 (7.8) | 0.286 |
| Heart failure | 20 (8.2) | 12 (8.5) | 8 (7.8) | 0.852 |
| Atrial fibrillation | 12 (4.9) | 9 (6.4) | 3 (2.9) | 0.369 |
| Chronic liver disease | 11 (4.5) | 5 (3.5) | 6 (5.9) | 0.387 |
| Stroke | 9 (3.7) | 7 (5.0) | 2 (2.0) | 0.310 |
| Gout | 6 (2.5) | 3 (2.1) | 3 (2.9) | 0.698 |
| Parkinson’s disease | 4 (1.6) | 2 (1.4) | 2 (2.0) | 1.00 |

Continuous data are presented as mean (standard deviation). Categorical data are shown as n (%). Comparison between participants with and without osteosarcopenia were conducted using Chi-square test or Fisher’s exact test for categorical variables, and Student’s t-test for continuous variables. ADL: Activities of Daily Living. IADL: Instrumental Activities of Daily Living. GERD: gastroesophageal reflux disease. MNA-SF: Mini Nutritional Assessment Short Form.

**Figure 1.**
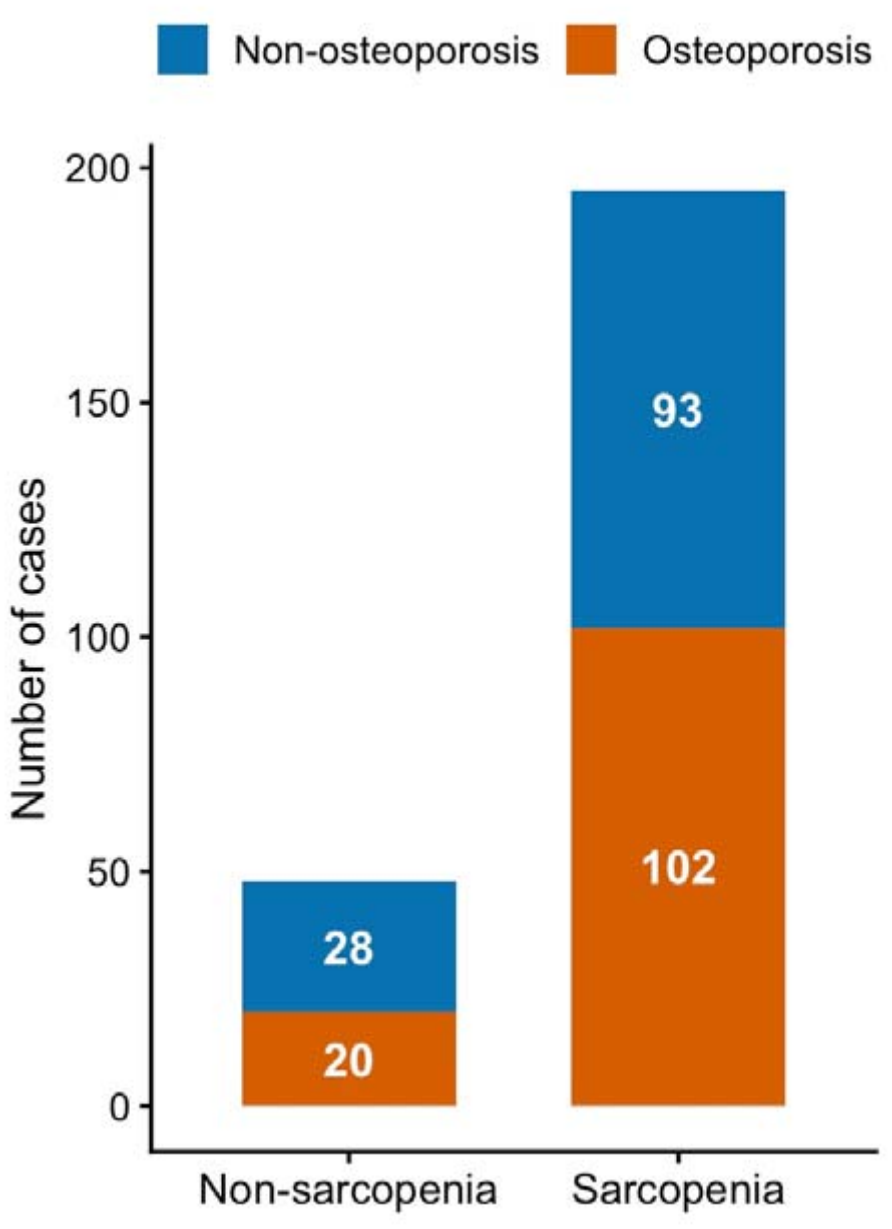
Number participants with sarcopenia, osteoporosis, and osteosarcopenia among 243 participants

The mean age of the participants was 75 years (SD 8. 6), and 65% were female. Participants with osteosarcopenia were significantly older than those without the condition (mean age 77/SD 8.0 years vs. 74/SD 8.7 years, p = 0.005). Female participants comprised most of the osteosarcopenia group compared to the non-osteosarcopenia group (84% vs. 50%, p < 0.001). Residential setting (rural versus urban) did not significantly differ between the groups. A higher proportion of participants with osteosarcopenia were underweight (20.6% vs. 12.8% in participants without osteosarcopenia), while a lower proportion were overweight (8.8% vs. 18.4% in participants without osteosarcopenia). The proportions of participants classified as having malnutrition or at risk of malnutrition were significantly higher among those with osteosarcopenia compared to those without osteosarcopenia.

In terms of comorbidities, hypertension (65.4%), gastroesophageal reflux disease (35.4%), and dementia (32.1%) were the most prevalent comorbidities across the entire study population. However, when comparing the two groups, coronary heart disease was the only comorbidity with a statistically significant difference, presenting more frequently in the osteosarcopenia group (35.3% vs. 22.7%, p = 0.031). All other assessed chronic conditions were similarly distributed between participants with and without osteosarcopenia.

### The factors associated with osteosarcopenia

**Table 2** presents the univariable and multivariable regression models of factors associated with osteosarcopenia. In the multivariable model, age (adjusted OR 1.05, 95%CI 1.01 – 1.09), female sex (adjusted OR 9.75, 95%CI 4.57 – 20.82), underweight (adjusted OR 2.48, 95%CI 1.03 – 5.96) were associated with higher odds of having osteosarcopenia. Participants with a BMI ≥25.0 had lower odds of having osteosarcopenia (adjusted OR 0.29, 95%CI 0.12 – 0.73) compared to those with BMI in the normal range.

**Table 2.** Univariable and multivariable logistic regression analysis of factors associated with osteosarcopenia.

| Factors | Unadjusted odd ratio (95% confidence interval) | p-values | Adjusted odd ratio (95% confidence interval) | p-values |
|---|---|---|---|---|
| Age (years) | 1.04 (1.01 – 1.08) | 0.006 | 1.05 (1.01 – 1.09) | 0.020 |
| Sex |  | <0.001 |  | <0.001 |
| Male (reference) | 1 |  | 1 |  |
| Female | 5.30 (2.83 – 9.93) |  | 9.75 (4.57 – 20.82) |  |
| Rural/urban living |  | 0.971 | - |  |
| Rural (reference) | 1 |  |  |  |
| Urban | 0.99 (0.43 – 2.24) |  |  |  |
| Body mass index (kg/m<sup>2</sup>) |  | 0.053 |  | 0.002 |
| Normal (18.5-24.9) (reference) | 1 |  | 1 |  |
| <18.5 | 1.57 (0.78 – 3.16) | 0.205 | 2.48 (1.03 – 5.96) | 0.042 |
| ≥ 25.0 | 0.47 (0.21 – 1.06) | 0.067 | 0.29 (0.12 – 0.73) | 0.008 |
| MNA-SF score |  | 0.005 |  | 0.182 |
| ≥ 12 (reference) | 1 |  | 1 |  |
| 8 – 11 | 3.26 (1.59 – 6.71) | 0.001 | 2.18 (0.95 – 4.97) | 0.065 |
| ≤7 | 2.94 (1.24 – 6.98) | 0.015 | 1.81 (0.65 – 5.00) | 0.252 |
| ADL impairment | 1.41 (0.83 – 2.41) | 0.209 | - |  |
| IADL impairment | 1.44 (0.77 – 2.70) | 0.253 | - |  |
| Comorbidities |  |  | - |  |
| Hypertension | 1.28 (0.75 – 2.19) | 0.373 | - |  |
| GERD | 1.07 (0.63 – 1.82) | 0.806 | - |  |
| Dementia | 1.19 (0.69 – 2.05) | 0.530 | - |  |
| Diabetes | 0.68 (0.39 – 1.18) | 0.170 | 0.62 (0.32 – 1.20) | 0.158 |
| Coronary heart disease | 1.86 (1.06 – 3.27) | 0.032 | 1.70 (0.87 – 3.32) | 0.119 |
| Osteoarthritis | 1.10 (0.58 – 2.07) | 0.781 | - |  |
| Dyslipidemia | 1.49 (0.78 – 2.85) | 0.222 | - |  |
| Chronic lung disease | 0.79 (0.38 – 1.65) | 0.532 | - |  |
| Chronic kidney disease | 0.73 (0.33 – 1.60) | 0.434 | - |  |
| Cancer | 0.62 (0.26 – 1.50) | 0.289 | - |  |
| Heart failure | 0.92 (0.36 – 2.33) | 0.852 | - |  |
| Atrial fibrillation | 0.44 (0.12 – 1.68) | 0.233 | - |  |
| Chronic liver disease | 1.70 (0.51 – 5.73) | 0.392 | - |  |
| Stroke | 0.38 (0.08 – 1.88) | 0.237 | - |  |
| Gout | 1.39 (0.28 – 7.05) | 0.688 | - |  |
| Parkinson’s disease | 1.39 (0.19 – 10.04) | 0.744 | - |  |

ADL: Activities of Daily Living. IADL: Instrumental Activities of Daily Living. GERD: gastroesophageal reflux disease. MNA-SF: Mini Nutritional Assessment Short Form. Variables with a p-value <0.20 on univariable analyses were included in the multivariable logistic regression model.

### The association between osteosarcopenia and falls during the 3-month follow up

Follow-up data were available for 208 participants, of whom 27 had a fall. The event rates for falls were 13.0% (27/208) overall, 19.8% (17/86) in the osteosarcopenia group vs. 8.2% (10/122) in the non-osteosarcopenia group (p = 0.014).

**Table 3** presents the unadjusted and adjusted odds ratios of osteosarcopenia on falls. In the unadjusted analyses, participants with osteosarcopenia had nearly three times the odds of experiencing a fall compared to those without the condition (OR 2.76, 95% CI 1.20 – 6.37). In the adjusted models, osteosarcopenia remained an independent predictor of incident falls after adjusting for age (adjusted OR 2.38, 95% CI 1.01 – 5.59), sex (adjusted OR 2.59, 95% CI 1.07 – 6.26), rural/urban location (adjusted OR 2.81, 95%CI 1.21 – 6.51), and the number of comorbidities (adjusted OR 2.71, 95% CI 1.17 – 6.30). When accounting for all candidate covariates (age, sex, rural/urban living, and number of comorbidities), there was still a positive association between osteosarcopenia and falls (adjusted OR 2.11, 95% CI 0.82 – 5.39), although not statistically significant.

**Table 3.** Odds ratios for falls comparing participants with and without osteosarcopenia.

|  | Falls |  |
|---|---|---|
|  | Odds ratios (95%CI) | p-values |
| Unadjusted | 2.76 (1.20 – 6.37) | 0.017 |
| Adjusted for age | 2.38 (1.01 – 5.59) | 0.047 |
| Adjusted for sex | 2.59 (1.07 – 6.26) | 0.035 |
| Adjusted for rural/urban living | 2.81 (1.21 – 6.51) | 0.016 |
| Adjusted for number of comorbidities | 2.71 (1.17 – 6.30) | 0.020 |
| Adjusted for sex, rural/urban living and comorbidities | 2.58 (1.05 – 6.36) | 0.039 |
| Adjusted for age, sex, rural/urban living and comorbidities | 2.11 (0.82 – 5.39) | 0.121 |

## Discussion

In this study of older hospitalised patients, 42% of the participants were found to have osteosarcopenia. Osteosarcopenia significantly increased the odds of falls in three months post-discharge. We found that older age, female sex, and underweight were associated with higher odds of having osteosarcopenia.

The prevalence of osteosarcopenia in our cohort (42.0%) appeared to exceed the pooled global and Asian community averages of 21–22%, which typically encompass broader populations aged 65 and older ^4^. However, this elevated rate is highly consistent with age-specific epidemiological trajectories when accounting for the advanced mean age of our study population (approximately 75 years). This finding closely aligns with data from the Bushehr Elderly Health (BEH) Program in Iran, which demonstrated that osteosarcopenia prevalence drastically increased with age, reaching 48.3% among women and 59.4% among men in the age group of 75 years and older ^22^. In another study of 76 older adults with a history of falls and hip fractures (mean age 81.0, SD 6.8 years) in Turkey, the prevalence of osteosarcopenia was 37% ^23^. Therefore, our cohort serves as a highly representative sample of this older, more vulnerable demographic.

Consistent with established global literature, our data also confirmed that the chance of having osteosarcopenia is significantly influenced by female sex, with female participants having nearly 10 times the odds of having osteosarcopenia compared to male participants ^4,5,24^. The predominance of this condition in females may be linked to post-menopausal hormonal changes. The sudden drop in estrogen levels can lead to increased bone resorption due to heightened osteoclast activity, as well as promotes muscle breakdown. This explains why older women, especially those in their mid-70s, are the most important demographic for early screening of osteosarcopenia^25,26^.

Our finding on the relationship between a low BMI (<18.50 kg/m^2^) and osteosarcopenia aligns with the established phenotype of sarcopenia and osteopenia in Asian populations^4,27,28^. Unlike Western demographics, where “sarcopenic obesity” is highly prevalent and excess fat masks muscle loss, osteosarcopenia in Asian cohorts is tightly linked to chronic protein-energy undernutrition and low body weight ^29,30^. According to the “mechanostat” theory, a lower body weight exerts less mechanical loading and gravitational stress on the skeletal and muscular systems, thereby failing to stimulate adequate bone mineralisation and muscle protein synthesis ^31^. Consequently, older Vietnamese patients presenting with low BMI should be immediately flagged as high-risk for dual musculoskeletal decline.

In this cohort, osteosarcopenia was identified as a strong, independent predictor of falls after 3 months. This finding aligns with a recent comprehensive analysis evaluating osteosarcopenia as a risk factor for falls ^32^. Although the association failed to reach significance after adjustment for several confounding factors, given the small number of events (27/208), including all five variables in the model would have increased the risk of overfitting, sparse-data bias, and inflated standard errors, as most established recommendations suggest the requirement of at least 10 events per variable ^33,34^.

The concurrent deterioration of skeletal muscle mass and bone architecture creates a state of severe mechanical instability. Sarcopenia directly impairs balance, postural control, and reactionary strength, while osteopenia compromises the structural integrity of the skeleton.^32^ Consequently, these patients lack both the neuromuscular control to prevent a stumble and the skeletal strength to absorb the impact, making falls a highly prevalent and immediate clinical consequence ^32^.

### Strengths and limitations

To the best of our understanding, this is the first study in Vietnam examining osteosarcopenia and its impact on falls in older hospitalised patients. However, the study has several limitations. First, it was conducted at one urban hospital, which may not be representative for all older patients in Vietnam. Second, the relatively short follow-up duration of three months may limit the ability to capture longer-term outcomes, such as readmission or mortality. In addition, odds ratios derived from logistic regression may overestimate associations for more common outcomes and should therefore be interpreted with caution.

## Conclusions

Osteosarcopenia was common among older participants in this study, and was associated with increased the odds of falls at three months post-discharge. Clinicians should consider screening for osteosarcopenia in older adults, particularly among women and those who are underweight. Further research is needed to examine the feasibility and effectiveness multidisciplinary prevention or rehabilitation programs for fall prevention for older adults with osteosarcopenia.

## Data Availability

The data that support the findings of this study are not openly available due to reasons of sensitivity and are available from the corresponding author upon reasonable request.

## Authors’ contributions

TVN, MTTBD and TNN conceived the study. TVN and MTTBD designed the study protocol, led the ethics application and conducted recruitment. TNN conducted the statistical analyses. TVN, MTD, and TNN wrote the manuscript. All authors were involved in data interpretation. The manuscript was revised for important scientific content by all authors. All authors approved the final version of the manuscript.

